# Enhanced Data Pre-processing for the Identification of Alzheimer’s Disease-Associated SNPs

**DOI:** 10.1101/2024.03.14.24303161

**Authors:** Juliana F Alves, Eduardo Costa, Alencar Xavier, Luiz Brito, Ricardo Cerri

## Abstract

Alzheimer’s Disease (AD) is a complex neurodegenerative disorder that has gained significant attention in scientific research, particularly since the Human Genome Project. Based on twin studies that utilize the resemblance of Alzheimer’s disease risk between pairs of twins, it has been found that the overall heritability of the disease is estimated at 0.58. When shared environmental factors are taken into account, the maximum heritability reaches 0.79. This suggests that approximately 58-79% of the susceptibility to late-onset Alzheimer’s disease can be attributed to genetic factors [4]. In 2022, it is estimated that AD will affect over 50 million people worldwide, and its economic burden exceeds a trillion US dollars per year. One promising approach is Genome-Wide Association Studies (GWAS), which allow the identification of genetic variants associated with AD susceptibility. Of particular interest are Single Nucleotide Polymorphisms (SNPs), which represent variations in a single nucleotide base in the DNA sequence. In this study, we investigated the association between SNPs and AD susceptibility by applying various quality control (QC) parameters during data pre-processing and rank the SNP associations through mixed linear models-based GWAS implemented in BLUPF90. Our findings indicate that the identified SNPs are located in regions already associated with Alzheimer’s Disease, including non-coding regions. We also investigated the impact of incorporating demographic data into our models. However, the results indicated that the inclusion of such data did not yield any benefits for the model. This study highlights the importance of GWAS in identifying potential genetic risk factors for AD and underscores the need for further research to gain a better understanding of the complex genetic mechanisms underlying this debilitating disease.

## 1. Introduction

### 1.1. Alzheimer’s Disease

Alzheimer’s Disease (AD) is a progressive neurodegenerative disorder that is primarily associated with aging. It is characterized by a range of cognitive and psychiatric symptoms that can lead to eventual disability [16]. As per projections by the World Alzheimer Report, the number of AD cases is expected to reach 131.5 million by 2050 [15]. Histopathologically, AD is characterized by synaptic damage and neuronal loss in brain regions responsible for cognitive function [16].

Given the significant worldwide impact of AD, researchers have aimed to identify genetic markers associated with the disease. The goal is to develop personalized medicine using data from these studies to improve the quality of life of individuals predisposed to AD.

In the mid-1990s, with the advent of sequencing DNA technologies, the Human Genome Project was initiated. It aimed to map and sequence the human genome, resulting in a significant increase in genetic data. The concept of personalized medicine has evolved to include tailored medical treatments based on an individual’s genetic features. Incorporating genetic risk factors, such as disease-associated Single Nucleotide Polymorphisms (SNPs), has improved disease prediction for individuals [6].

There are three primary genomic approaches employed by researchers to explore AD. The first approach is Genetic Linkage Analysis, which aims to identify chromosomal regions associated with the disease without relating them to a specific gene [17]. The second approach involves the study of candidate genes, which compares the genetic variation between healthy individuals and AD patients. This approach has identified alleles in the APOE gene as strong candidates for AD [11]. The third approach is Genome-Wide Association Studies (GWAS), which aims to identify SNPs associated with increased susceptibility to AD.

### 1.2. GWAS

The focus of this study is to identify SNPs that are associated with AD, using genome-wide association studies. SNPs, or Single Nucleotide Polymorphisms, refer to variations at specific locations in the DNA chain. These variations are categorized based on the type of nucleotide substitution that occurs. For example, substitutions of C→T or G→A are referred to as transition SNPs, while substitutions of C→A, A→T, T→C, or T→G are referred to as transversion SNPs [3].

SNPs are the most commonly occurring type of genetic polymorphism in human genetics, and can impact various human traits and their development in a specific environment. Additionally, SNPs are evolutionarily stable, meaning that there is little variation among different generations [3].

The workflow of a GWAS involves multiple steps that are carefully planned to avoid biases and errors. The first step is to obtain data from a comprehensive and diverse group of individuals, which includes genotypic and phenotypic information for both the case and control populations. Genotypic information corresponds to the DNA obtained using SNP arrays or sequencing strategies. Before proceeding to the association test, it is recommended to apply a quality control procedure to the data to increase its reliability. This involves performing data cleaning using statistical approaches and biological concepts to mitigate biases that could interfere with the results. In this study, we emphasize the impact of Quality Control on GWAS by creating multiple scenarios, as explained in Subsection 1.3. Following the association test, multiple post-GWAS analyses can be conducted to interpret the results.

### 1.3. Data Processing for Quality Control

The input data for the association analysis includes individual ID numbers, disease stage, sex, and SNPs obtained through sequencing, along with information about the genotype batch. To minimize errors and bias, Quality Control (QC) techniques are required for the input data. These methods filter SNPs and individuals using statistical and mathematical equations, based on biological concepts.

To preserve data quality control and significantly reduce the number of candidate SNPs, several statistical filters are applied in GWAS [1]. These techniques prevent the effects of SNPs from being masked by the high dimensionality of the dataset. In this study, we present traditional methods used in GWAS to filter SNPs, to avoid the situations mentioned above. The filters used in our study are as follows:

- **Minor Allele Frequency (MAF):** In population genetics, the term “minor allele frequency” (MAF) refers to the frequency of a less prevalent allele at a particular locus (position) in a population. It is described as the prevalence of the minor allele, or second most frequent allele, at a specific locus in a population. The major allele, which is the more prevalent allele at that locus, is opposed by the minor gene. In GWAS, the calculated value for MAF can decrease the number of SNPs by deleting rare variants from the database. Hence, MAF is the frequency of the second most common allele in the studied population. SNPs with MAF less than a specified threshold (e.g. 1%) are removed [19];
- **Linkage Disequilibrium (LD):** The non-random association of alleles at various loci in a community is known as linkage disequilibrium (LD). In other words, it refers to the propensity of specific alleles to be inherited together more frequently than would be predicted by chance at various loci. In GWAS, researchers usually genotype a significant number of SNPs throughout the genome in cases and controls to find SNPs that are connected to the illness or trait under study. Researchers can find clusters of SNPs that are in LD and therefore likely to be passed together by looking at the patterns of LD between SNPs. These SNP clusters are known as haplotypes, and they can be used to locate parts of the genome that are linked to a particular illness or trait. Therefore, the use of LD as a filter serves to guarantee quality control and to prevent information loss [9];
- **Hardy-Weinberg Equilibrium (HWE):** Hardy-Weinberg equilibrium assumes that in a Mendelian population allele frequencies will remain constant across generations. Thus it is used to remove variants that do not conform to its expectations;
- **Genotype Missingness (GENO) and Sample Missingness (SAMPLE):** For these both parameters the filtering procedure is applied by choosing a threshold and calculating the missingness of either Genotype or Sample data. Missingness above the threshold is deleted from the database.

Investigating the population structure in a Genome-Wide Association Study (GWAS) is crucial, as it can result in misleading outcomes when predicting and selecting genetic variations that depict the issue at hand [8]. Population stratification refers to differences in the frequency of genetic variants between groups of cases and controls due to systematic ancestral differences [19]. In this work, the phenotype of interest is the occurrence of Alzheimer’s Disease, and we conducted an Analysis of Variance (ANOVA) to investigate whether the population structure would affect the model.

## 2. Methods

In this section, we describe the data utilized in this study and its acquisition. Additionally, we elucidate the data processing pipeline, encompassing the data transformation, cleaning, and validation steps. Finally, we illustrate how demographic information such as gender, race, ethnicity, and age of the individuals were obtained to understand their influence on the phenotype.

### 2.1. Data

The data used in this study was obtained from the Alzheimer’s Disease Neuroimaging Initiative (ADNI) ^1^ database. Launched in 2004 as a public-private partnership, ADNI aims to identify whether brain images, biological markers, and clinical and neuropsychological assessments can be combined to measure the progression of mild cognitive impairment (MCI) and early Alzheimer’s disease (AD).

The genotype data initially contained 620,901 variants, with a mean missingness of 30,785 variants. The input file included 757 individuals, comprising 449 men and 308 women. The phenotype was divided into three categories: Normal (CN), Alzheimer’s Disease (AD), and Mild Cognitive Impairment (MCI). These categories were estimated by ADNI using various biomarkers, which are substances, measurements, or indicators of a biological state that may be identified before clinical symptoms appear.

### 2.2. Data Processing

The data processing pipeline included several steps for data transformation, cleaning, and validation, as well as demographic information extraction, including gender, race, ethnicity, and age. The impact of this information on the phenotype was also investigated in this study.

#### 2.2.1. Data Manipulation

The initial dataset consisted of three separate files: a genotype file (Table 2), a map file for identifying SNP ids in the genotype file (Table 3), and a phenotype file (Table 4). To consolidate this information into a single file, we utilized the PLINK software, a whole-genome association analysis toolset. With PLINK, we successfully merged the separate files and applied the filters mentioned in Subsection 1.3 to create the final dataset file (Table 1). This consolidated file is suitable for use in GWAS analyses and Machine Learning models.

**Table 1:** Dataset template layout: PLINK output file.

| Individual ID | Phenotype | rs3094315 | rs12563034 | ... | MitoC16272T |
| --- | --- | --- | --- | --- | --- |
| subject1 | CN | 0 | 1 | ... | 0 |
| subject2 | AD | 0 | 0 | ... | 1 |
| subject3 | CN | 2 | 2 | ... | 2 |
| ... | ... | ... | ... | ... | ... |
| subject_n | MCI | 1 | 1 | ... | 2 |

**Note:** This file maps genotypes and health diagnoses for each individual. SNP information is presented in columns, with each column representing an allele. SNP names are obtained from the MAP file.

**Table 2:** PLINK genotype input file.

| Individual ID | Genotype | **Note:** Column 1: Individual ID; Column 2: Genotyping data, where 0 represents homozygous, and 1 and 2 represent different heterozygous types. |
| --- | --- | --- |
| subject1 | 0102122102120 |  |
| subject2 | 0112120212012 |  |
| ... | ... |  |
| subject_n | 0212120211011 |  |

**Table 3:** PLINK map input file.

| SNP ID | Chromosome | Position |
| --- | --- | --- |
| rs3094315 | 1 | 742429 |
| rs12563034 | 1 | 758311 |
| ... | ... | ... |
| MitoC16272T | 26 | 16272 |

**Note:** The purpose of the MAP file is to identify the location of each SNP within the chromosome, as well as its corresponding SNP ID. The order of the SNP IDs in the MAP file matches the order of the genotype data in Table 1, column 2.

**Table 4:** PLINK phenotype input file.

| Individual ID | Phenotype | **Note:** This file provides a mapping of the health diagnoses for each individual, where a value of 1 indicates cognitive normalcy, a value of 2 indicates mild cognitive impairment, and a value of 3 indicates Alzheimer’s disease. |
| --- | --- | --- |
| subject1 | 1 |  |
| subject2 | 3 |  |
| subject3 | 1 |  |
| ... | ... |  |
| subject_n | 2 |  |

#### 2.2.2. Samples demographics analyzes

In addition, during this study, the effects of demographics on the phenotype were analyzed to verify if they would be used as parameters in the models. The demographics include information about the individuals such as race, gender, ethnicity, and age. If relevant, these data would be used to train the models and improve the SNPs selection. To analyze the impact of these features, a linear model was used to calculate the Analysis of Variance (ANOVA) and obtain the feature importance of each demographic variable. ANOVA is a statistical test that determines whether the observed correlations in a sample can be generalized to the entire population. Therefore, the results from this test indicate whether it is reasonable to draw any conclusions based on the sample and if it is valid to use certain demographic information to train the model. In light of this, prior to incorporating demographic variables into the models, we examined their significance through ANOVA.

#### 2.2.3. Quality Control

In this section, we discuss how statistical filters were applied to guarantee quality control on the dataset. Quality control (QC) is a step in data pre-processing, and it improved data consistency and validity of the results. Thus, we created multiple datasets by varying the values for each QC hyperparameter during the QC stage, as shown in Table 5.

**Table 5:**
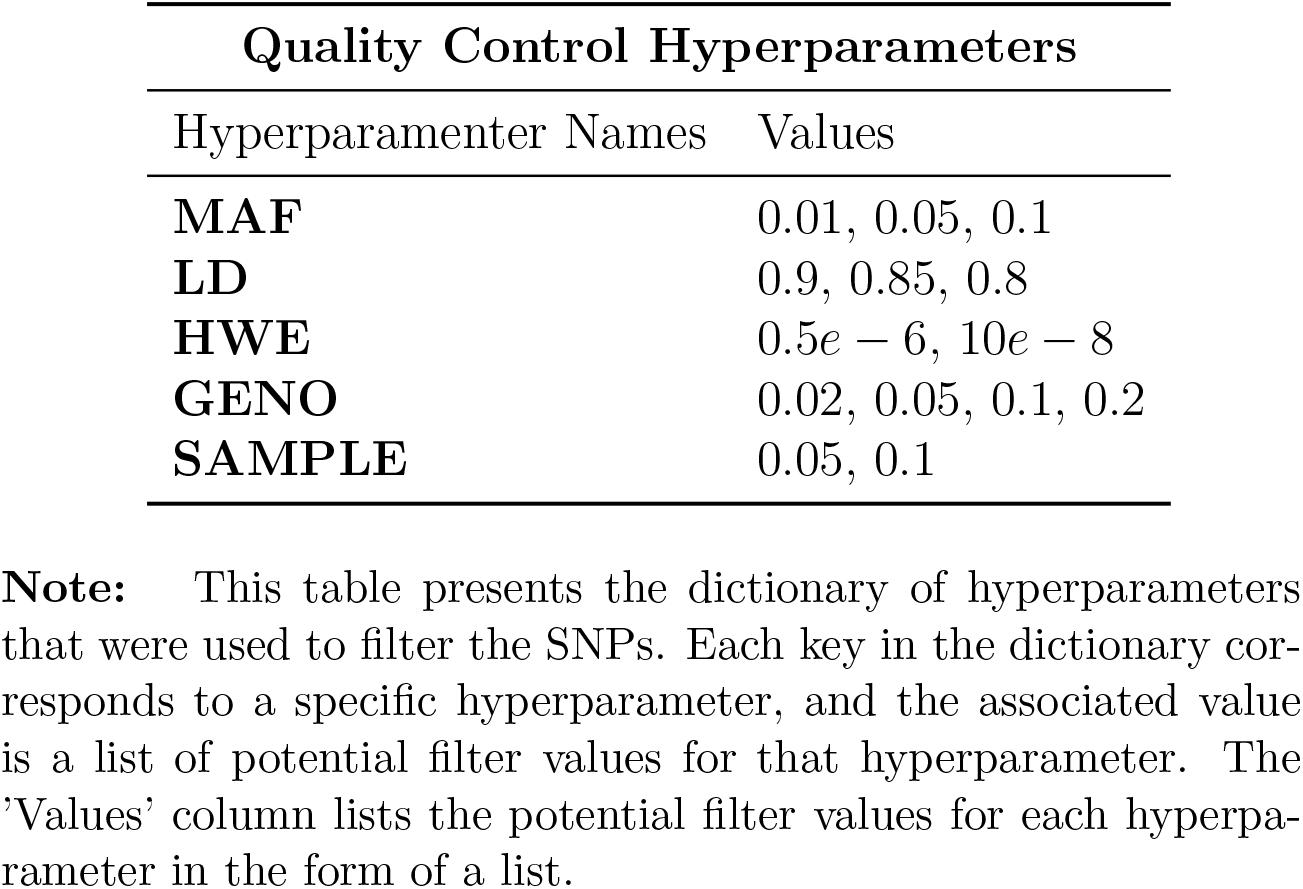
Parameters to guarantee quality control.

Each combination of quality control parameters resulted in a distinct dataset that was then used to generate a statistical model. Because each set of quality control parameters produced a unique dataset and subsequent model, we refer to each iteration as a “round”. In other words, a round is a specific combination of QC hyperparameters that resulted in a particular dataset, which was then used to build a model. By conducting multiple rounds with varying combinations of quality control parameters, we could explore the impact of these parameters on the final model’s performance and identify the optimal set of parameters for our analysis.

To illustrate, consider the 1st round, which consists of a dataset generated by combining QC hyperparameters. Specifically, the dataset has a genotype missingness rate of 0.02, a sample missingness rate of 0.05, a minor allele frequency (MAF) of 0.01, a Hardy-Weinberg equilibrium (HWE) threshold of 5e-6, and a linkage disequilibrium (LD) cutoff of 80.

In this study, we performed QC using various parameter combinations, and the resulting datasets were compared to assess the impact of each parameter on the data quality. We evaluated the quality of the datasets using different metrics, such as the percentage of missing data, the number of samples and SNPs after QC, and the deviation from the expected Hardy-Weinberg equilibrium. Subsequently, we assessed the impact of QC on downstream analyses, such as principal component analysis and genome-wide association analysis.

The filters listed in Table 5 were successfully applied to the data using the PLINK toolset and a Python script that automated the process, resulting in the creation of 144 filtered data files. Each of these data files was subjected to GWAS analyses in this study.

To use PLINK, the data must be formatted into one of several standard acceptable input formats, including .vcf, binary, or text files. The ADNI dataset was pre-formatted in binary format.

### 2.3. GWAS and genomic prediction

The BLUPF90 is a family of programs designed for the computation of mixed linear model with a focus on breeding applications. This software was utilized for GWAS and fitting the genomic model in this study. BLUPF90 offers a set of functionalities, including data QC, variance estimation using various methods, the estimation of Best Linear Unbiased Estimators (BLUEs) and Predictors (BLUPs) for large datasets, computation of individual-level accuracy, allows for the utilization of pedigree and SNP information to estimate genetic merit and conduct genome-wide association studies (GWAS) [10]. BLUPF90 program was developed to analyze large datasets for breeding applications at high-performance and without need for coding.

This study focuses on using a subset of programs from the BLUP family of programs, being them: RUNUM, THRGIBBS1F90, POSTGIBBf90, BLUPf90, and POSTgs. We built a pipeline using these programs, as shown in Figure 1. Following is a description of each of the applications that were used.

**Figure 1:**
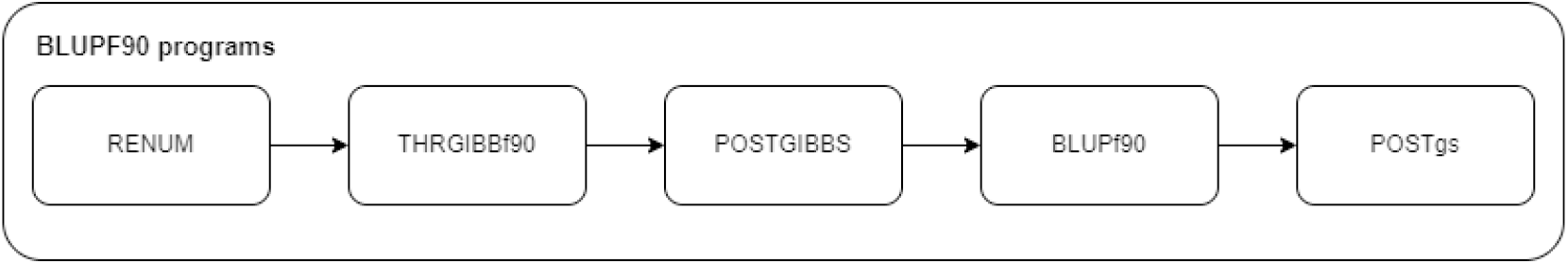
Execution pipeline of the BLUPF90 programs

1. RUNUM is used to generate the parameter file (runf90.par) as shown in Figure 2. A parameter file includes the statistical model, parameters and options to run the programs in the pipeline. Parameters and options are described in the BLUPF90 manual ^2^.
2. THRGIBBf90 runs the Markov chain Monte Carlo samples for the estimation of variance components of the Mixed Linear Models (MLM) of a threshold model, utilized for categorical response variables. MLM differ from general linear models (GLM) by modeling random effect terms other than the residual term. Thus, the random component *ε*, with variance *V* [*ε*] = *R*, of GLM (Equation 1) is extender to *ε* = *Zu*+*e*, with variance *V* [*ε*] = *ZGZ*^*′*^ + *R*, as the variance of the random terms are *V* [*Zu*] = *ZGZ*^*′*^ and 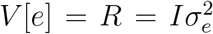, so that 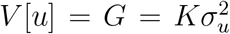. Thus, the variance component estimation is a key operation of MLMs.

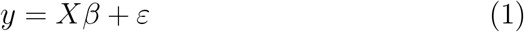

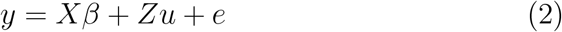

This method uses Gibbs Sampling (GS) for the MCMC sampling of variance components. GS samples each individual parameter from their univariate full-conditional distribution as an efficient way to sample a multivariate probability *p*(*X, Y*) [2].
3. POSTGIBBSF90 summarizes the MCMC samples to generate the posterior mean estimators of the variance components (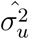 and 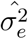.
4. BLUPF90 uses the variance components from POSTGIBBSF90 to fit the genomic model (2) to infer the genetic merit of individuals. Herein, this corresponds to the probability of individuals to fall into one of three phenotypic categories (i.e., CN, MCI and AD).
5. POSTgs is a post hoc procedure to extracts SNP solutions from the genetic merits, as shown in 3. Subsequently, it computes p-values for all SNPs. The significance of these associations allows for comparison of the impact of data quality control for the various scenarios under consideration.

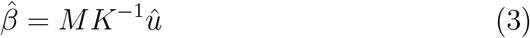

The p-value is the probability of the estimated deregressed SNP effect (*α*) given the the null-hypothesis (*H*_0_) is true, thus *p*(*α*_*j*_|*H*_0_). For the *j*^*th*^ SNP, this is computed according to the Equation 4.

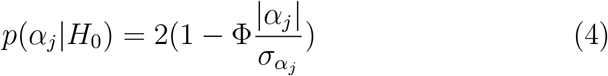

The deregressed SNP effects (*α*) and their standard deviation are estimated from *β* rescaled by diagonal elements of the left-hand side equation utilized to solve the mixed model equation. The equations are described by Aguilar et al. (2019).

**Figure 2:**
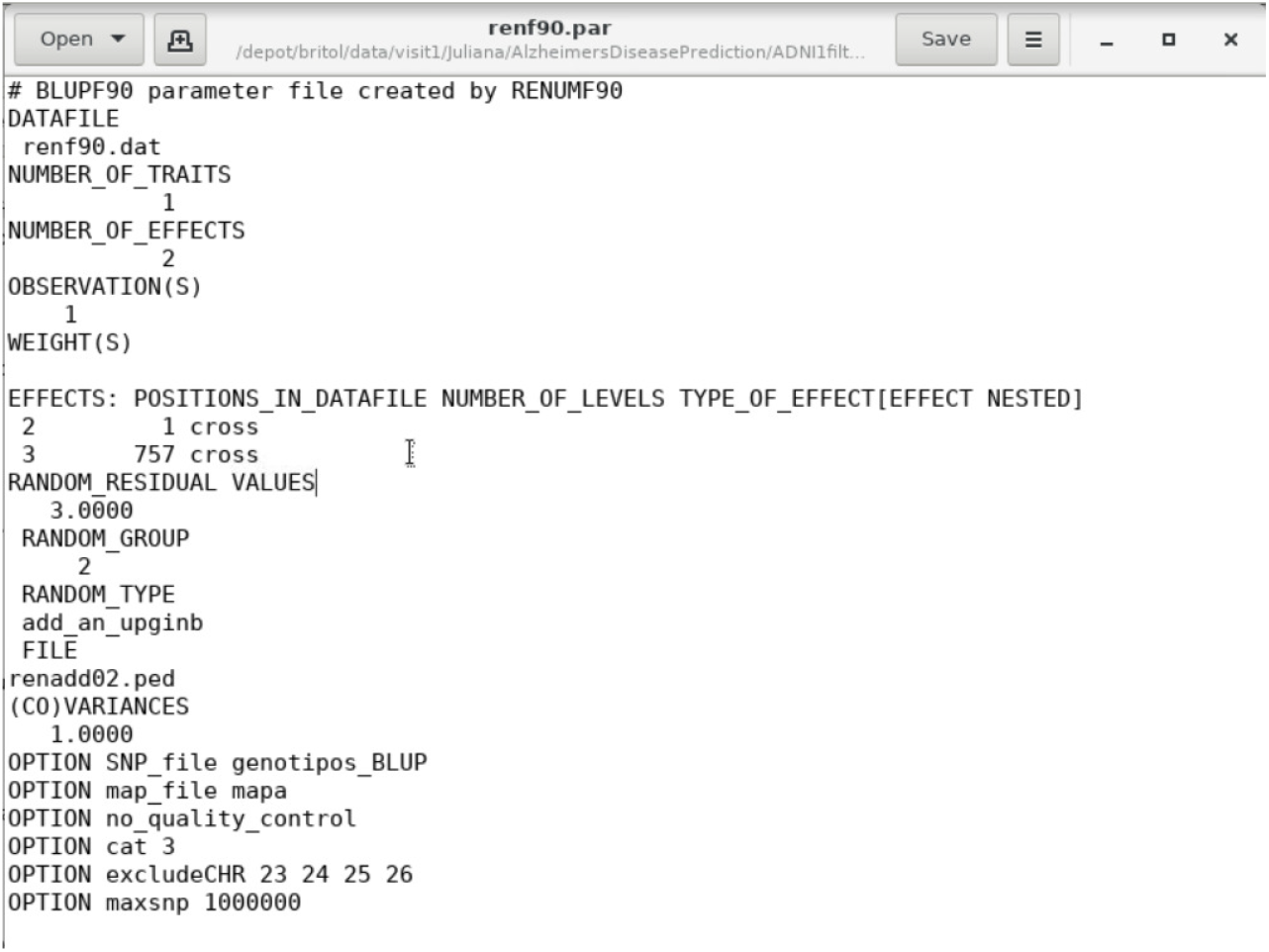
Parameters card (par file) used to specify the parameters and options to run the BLUP family of programs

## 3. Results and Discussion

In this section, we describe the results obtained from this study. As explained in Subsection 2.2.3, quality control was applied to data by statistical filters. Later, we used the BLUP Family of Programs to model the problem and find the most significant SNPs for the Alzheimer’s Disease phenotype. Furthermore, alongside the quality control phase, we analyzed the individual demographic data to determine their suitability for use in the models. Additionally, we compared the SNPs with the most significance to those reported in the literature to be associated with AD. SNPs with higher significance may provide valuable information for studying Alzheimer’s Disease cases.

### 3.1. Demographics understanding

The ANOVA method was used to comprehend the significance of using Age, Race, Ethnic, and Gender on the models to predict Alzheimer’s Disease. The first hypothesis was that at least age would be associated with the disease. The ANOVA results are shown in Table 6. In the results obtained, the p-values are always much greater than 0.05, for this reason, they are not significant input in the model to assume someone’s likelihood to have Alzheimer’s Disease.

**Table 6:** ANOVA: demographics data.

|  | Degrees of freedom | P-value |
| --- | --- | --- |
| **Gender** | 1.0 | 0.87390 |
| **Race** | 4.0 | 0.35020 |
| **Ethnic** | 2.0 | 0.60523 |
| **Age** | 1.0 | 0.47378 |

### 3.2. Data Quality Control

Here we highlight the impact of varying the statistical filters for quality control. In this manner, the datasets suffered cut-offs that will be exposed along variations of significance values in each round.

Among the datasets generated, the most favorable outcome was observed in the dataset characterized by a minor allele frequency (MAF) of 0.01, a linkage disequilibrium (LD) threshold of 85, a Hardy-Weinberg equilibrium (HWE) significance of 5e-6, a sample missingness rate of 0.01, and a gene missingness rate of 0.1. Notably, the most significant result was obtained for the SNP rs7918269, with a p-value of 1.600000e-09. This p-value was over 10 times larger than the second-best value encountered. The following images show the impact of varying the QC hyperparameters and how the best result encountered could be sweet spot.

**Figure 3:**
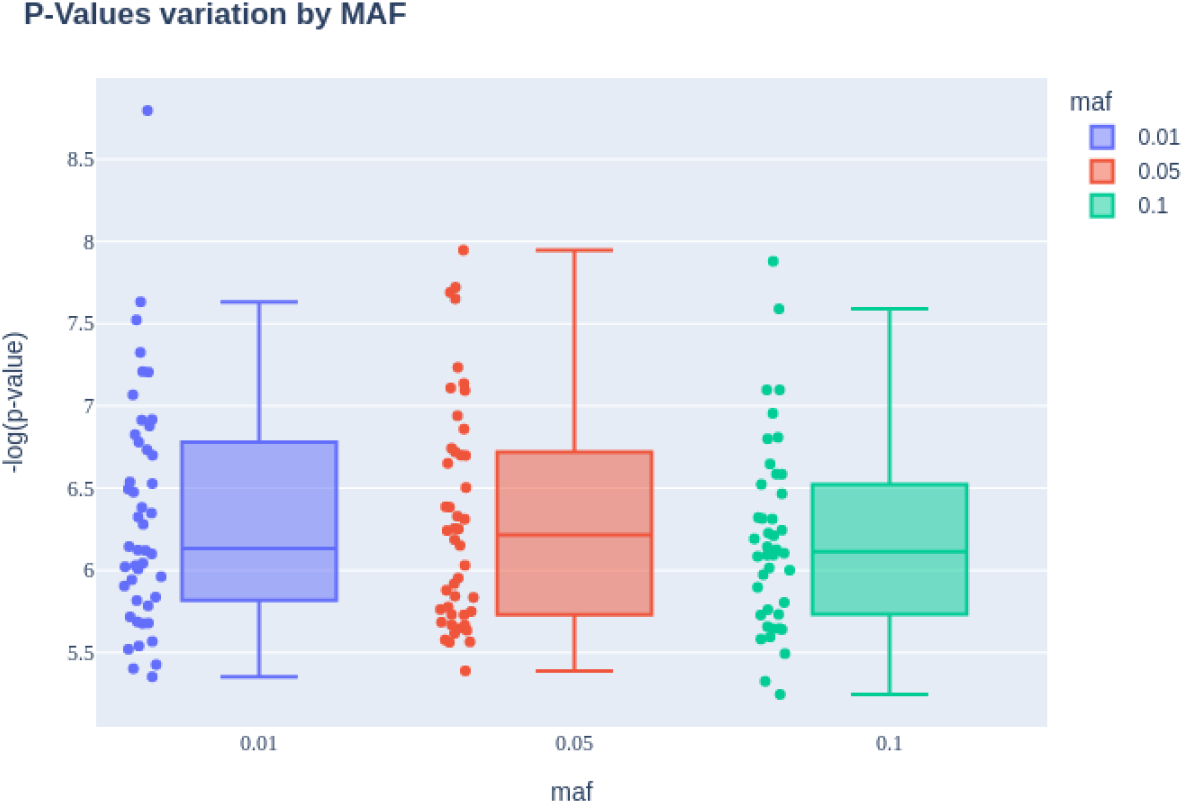
P-values variation by Minor Allele Frequency

**Figure 4:**
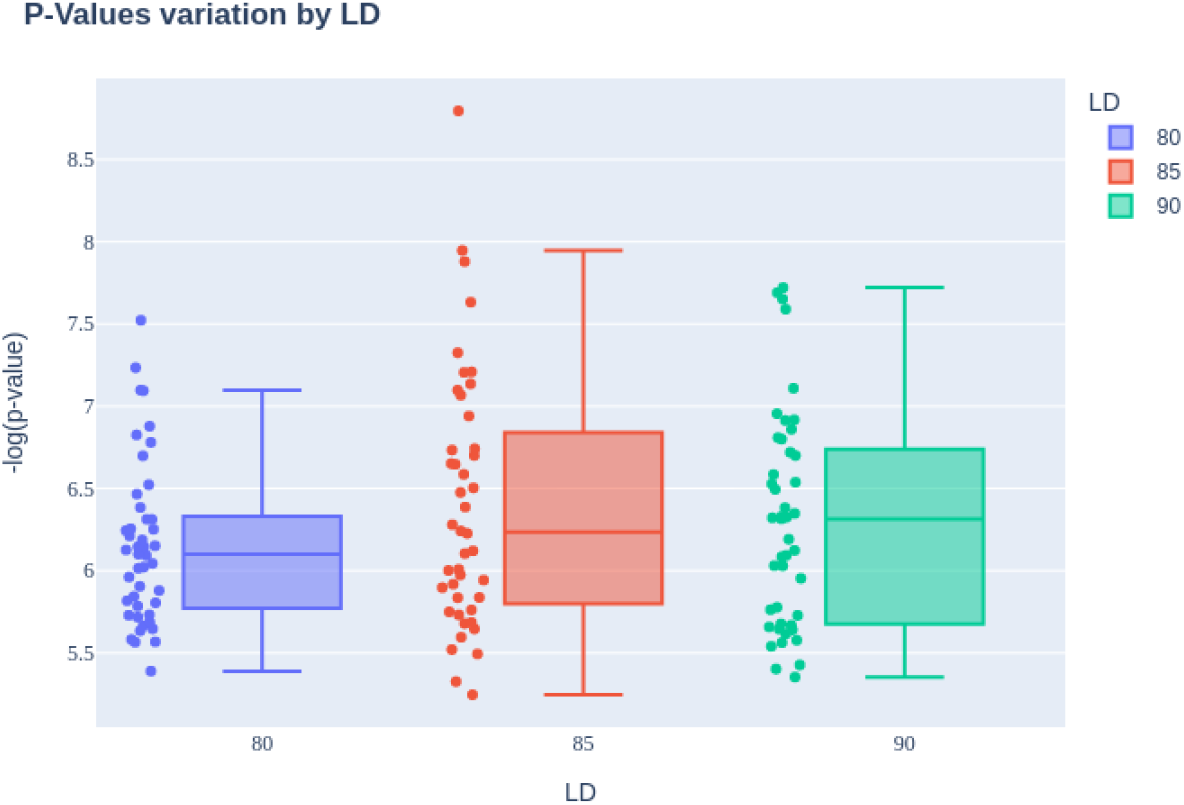
P-Values variation by Linkage Disequilibrium

**Figure 5:**
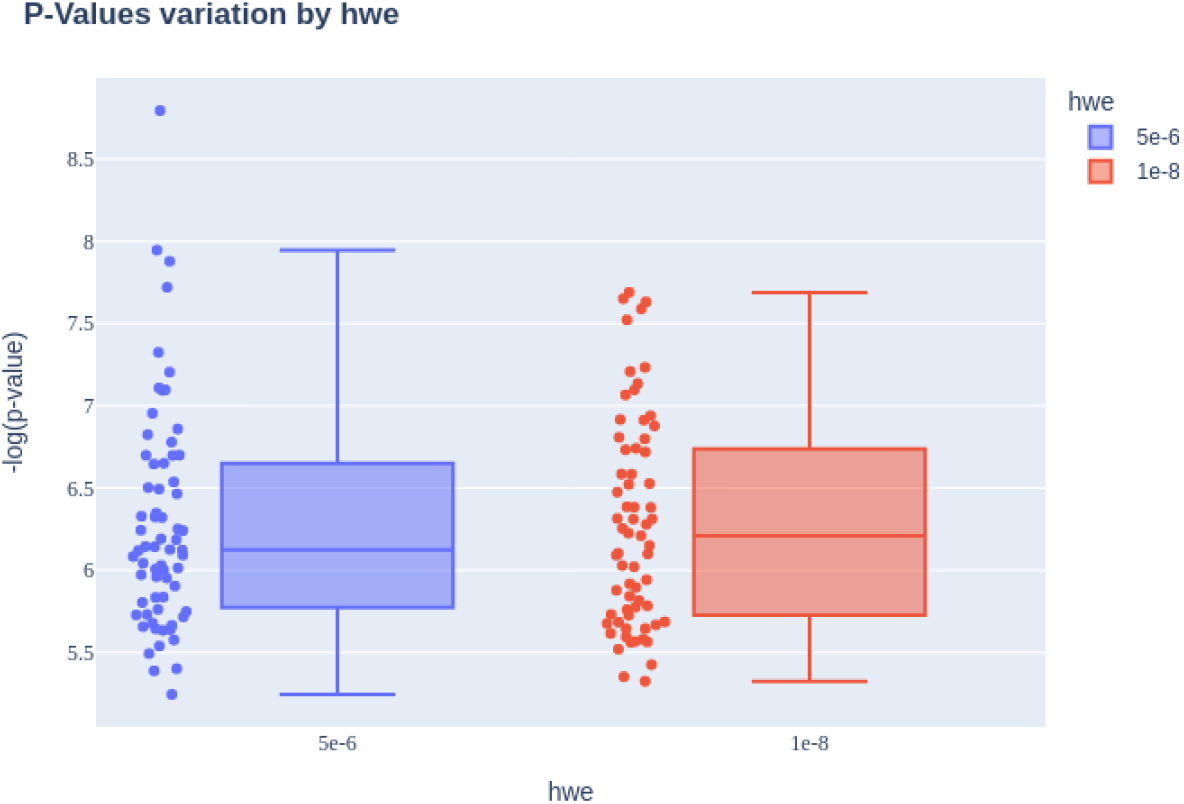
P-Values variation by Hardy-Weinberg equilibrium (Created by author)

**Figure 6:**
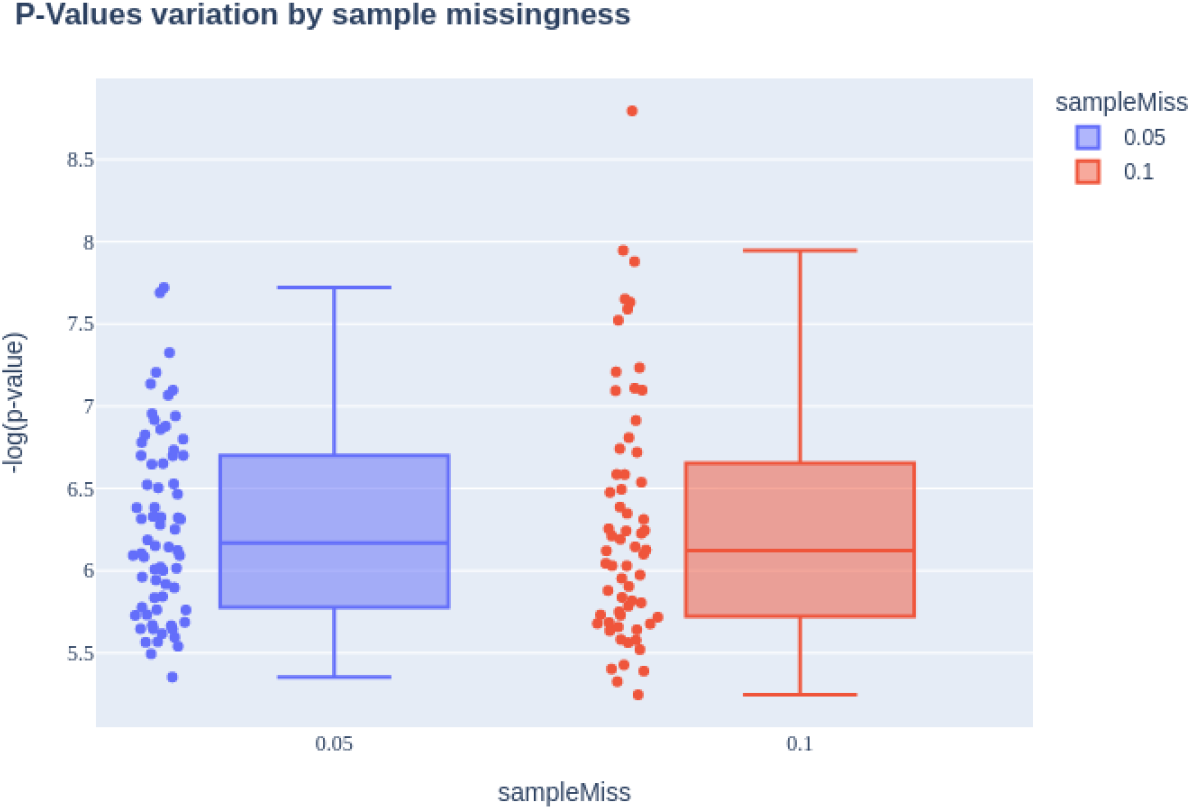
P-Values variation by Sample Missingness (Created by author

**Figure 7:**
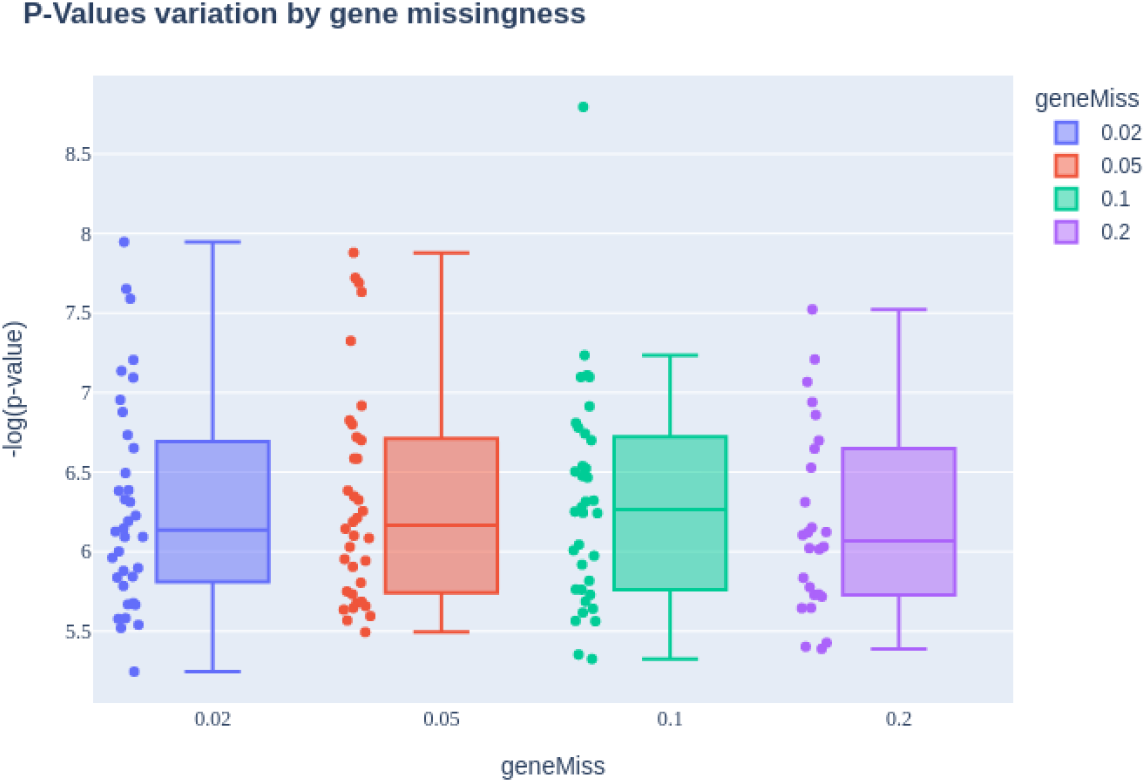
P-Values variation by gene Missingness (Created by author

### 3.3. SNPs selection using BLUPF90

To analyze and evaluate each round, we created a table containing the 1000 most significant SNPs from each round. As shown in Table 7, for each round there are two columns. The first column represents the SNP Name and the second column represents its p-value. In this manner, each round is ordered from the lowest to the highest p-value.

**Table 7:** Illustration of results: Ranked SNP p-values for each set of parameters.

|  | round 1 |  | round 2 |  | ... | round 144 |  |
| --- | --- | --- | --- | --- | --- | --- | --- |
|  | SNP Name | SNP P-value | SNP Name | SNP P-value |  | SNP Name | SNP P-value |
| 0 | rs17184445 | 0.000001 | rs738413 | 6.2300e-8 | ... | rs7158356 | 4.881000e-07 |
| 1 | rs713158 | 0.000004 | rs17107651 | 7.3900e-7 | ... | rs7863479 | 4.131600e-06 |
| 2 | rs7914151 | 0.000005 | rs1448333 | 1.5356e-6 | ... | rs2027767 | 4.601000e-06 |
| 3 | rs608862 | 0.000006 | rs9681884 | 5.5215e-6 | ... | rs10495181 | 4.627100e-06 |
| 4 | rs12946426 | 0.000001 | rs9941427 | 8.7564e-6 | ... | rs10894598 | 5.116100e-06 |
| ... | ... | ... | ... | ... | ... | ... | ... |
| 999 | rs2617671 | 0.001010 | rs10867501 | 9.755883e-04 | ... | rs4965091 | 1.327353e-03 |

This representation for the rounds simplifies the access to each round of data. Besides that, it will help to answer questions such as “what round has the highest SNP value?”, “Does the SNP with higher significance appears in high positions in other rounds?”, and “Which SNPs appear more times in the top-ranked SNPs with significance value in all rounds?”.

Firstly we selected the 20 most significant SNPs in the whole table. In other words, all SNPs in the table were sorted from the lowest to the highest p-value, but only the 20 SNPs with the lowest p-values were selected. After that, we were able to map those SNPs to their respective gene by using the NCBI search engine.

After searching their position in the genome and getting their respective gene, we found 8 genes already associated with Alzheimer’s Disease in other studies. These genes are RUNX2-AS1, CNTN6, KLF12, RPS6KA2, MLN, FANCC, RUNX2, and ADCY9.

Once the location in the genome of each of the 20 SNP was found, we compared them with results from other studies. Table 8 shows this ranking and which gene the SNP belongs to. Some SNPs belong to uncharacterized genes, thus this work compared only SNPs in coding regions.

**Table 8:** SNPs with highest significance value among the rounds.

| SNP Name | Gene |
| --- | --- |
| rs105375077 | RUNX2-AS1 |
| rs105375024 | LOC105375024 |
| rs105374660 | LOC105374660 |
| rs105370220 | LOC105370220 |
| rs102723906 | LOC102723906 |
| rs100887750 | MRPS31P5 |
| rs27255 | CNTN6 |
| rs11278 | KLF12 |
| rs6196 | RPS6KA2 |
| rs4295 | MLN |
| rs2176 | FANCC |
| rs860 | RUNX2 |
| rs115 | ADCY9 |
| rs105378215 | CTB-99A3.1 |
| rs105378178 | LOC105378178 |
| rs100887750 | MRPS31P5 |
| rs57628 | DPP10 |
| rs54877 | ZCCHC2 |
| rs27255 | CNTN6 |
| rs23544 | SEZ6L |

RUNX2-AS1 was related to AD due to its impact on anxiety-like behavior [7], and was categorized as a novel biomarker for AD [18]. In [14] CNTN6 is described as a gene that modulates hippocampal synaptic plasticity and behavior, and was recognized as a risk gene for neuropsychiatric disorders. KLF12 is recognized as the gene with the second highest importance when analyzing AD-associated genes from blood [13]. RPS6KA2 is a gene involved in Neurotrophin signaling, and has had previous reports of association with Parkinson’s Disease in GWAS studies [12]. FANCC has been associated with entorhinal cortex thickness, a region that is involved early in the development of Alzheimer’s disease [5].

In the same context, we wanted to obtain which SNPs appear in the best positions in the rounds most of the time. By doing it we can analyze if significant SNPs in most rounds are in genes related to AD, and also compare them with future results from Machine Learning models. Table 9 shows the 10 SNPs that appeared in most of the rounds. As can be seen, there are three SNPs in genes, which are protein-coding regions. Therefore, a bibliographic research was performed about the genes ZFAND4, NCAM2, and LOC105372740 with the goal of finding any relation to Alzheimer’s Disease. In the bibliographic research, it was found that ZFAND4 protein domains are involved in regulating the immune response when interacting with other genes. Genetic data have implicated the NCAM2 gene in neurodevelopmental disorders including Down Syndrome, autism, and Alzheimer’s Disease.

**Table 9:**
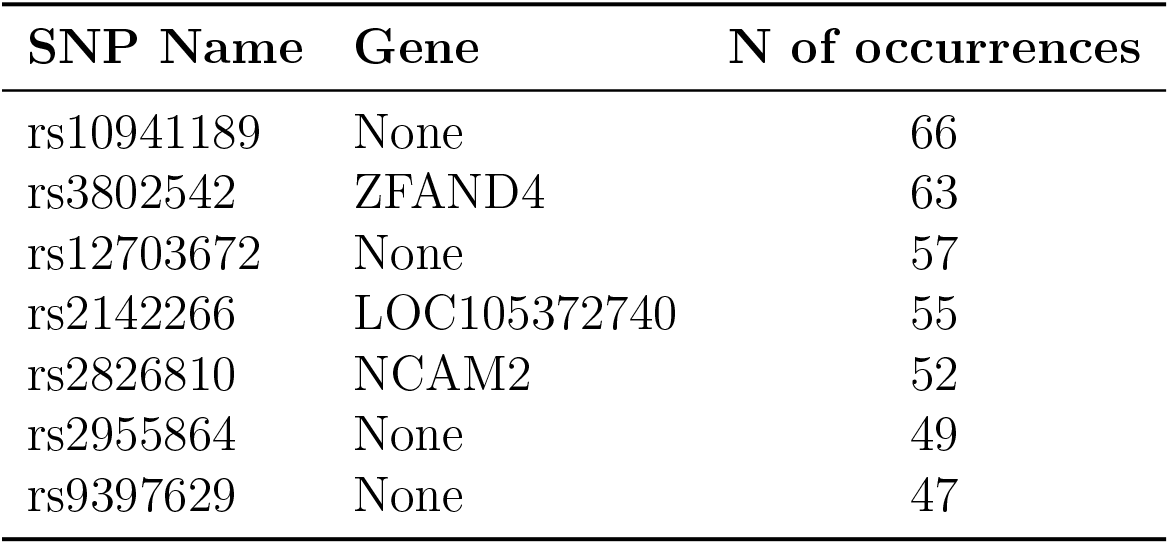
SNPs with most occurrences.

## 4. Conclusions and Future Work

In this study, we have developed a pipeline to compare the impact of various parameters on Quality Control, namely MAF, GENO, MIND, HWE, and LD. The implementation of this pipeline has provided a deeper understanding of genomic tools for Genome-Wide Association Studies (GWAS) and Quality Control pruning, specifically using PLINK and BLUPF90. By employing genomic selection with the BLUPF90 family of programs, we have identified candidate genes for Alzheimer’s Disease. Some of these genes are already known to be associated with Alzheimer’s Disease in the literature, while others represent new findings.

Demographic information did not prove to be significant predictors for estimating the desired phenotype, as determined by ANOVA, thus was not accounted for in the genomic model.

Moving forward, we plan to employ Machine Learning (ML) models on the best Quality Control round. We will select the 600 SNPs from the best round and use k-fold cross-validation (k=5) to train our models. Specifically, we will use THRGIBBS to calculate Genomic Best Linear Unbiased Predictions (GBLUPs) and Random Forests to determine SNP’s significance, while avoiding interference with the prediction process. Subsequently, we will use GBLUPs to predict the phenotype, which will serve as a rough replacement for LASSO Linear Regression. Finally, we will use Random Forest to predict Alzheimer’s Disease using the remaining 20% of the data.

## Data Availability

All data produced are available online at https://adni.loni.usc.edu/

## Footnotes

1 http://adni.loni.usc.edu

2 http://nce.ads.uga.edu/wiki/lib/exe/fetch.phpmedia=blupf90_all8.pdf

